# Removing Redundant Anatomical Inputs Improves Deep Learning-Based Forensic Dental Age and Sex Estimation

**DOI:** 10.64898/2026.10.05.26364771

**Authors:** Zongze Wu, Weijie Shan, Wujun Zhang, Zhenpei Kang, Benyu Yang, Zibin Liao, Zehang Huang, Yan Zhang, Xia Yue

**Author notes:** Corresponding authors: X.Yue; Y.Zhang. These authors contributed equally to this work (Zongze Wu, Weijie Shan). These authors contributed equally to this work (Wujun Zhang, Zhenpei Kang, Benyu Yang, Zibin Liao, Zehang Huang).

## Abstract

Age and sex estimation from dental radiographs supports forensic identification and age-dependent legal assessment. Multi-input models may combine global and regional representations, but when these inputs are derived from the same radiograph, they may repeat anatomical information rather than provide complementary signals. We developed HMA-Net, a hierarchical multi-stream anatomical network, together with a task-specific input audit that evaluates whether each candidate representation improves at least one prediction task without materially degrading the other. The model was developed using a retrospective single-centre cohort of 4,293 panoramic radiographs from individuals aged 3–18 years and combined a panoramic-image stream with graph-linked tooth representations. On the held-out test set, the final panoramic-plus-permanent-tooth model achieved a mean absolute age error of 0.51 years and a sex-classification accuracy of 85.2%; 99.4% of age estimates were within 2 years of chronological age. Across three independent training runs, removing the jaw-region stream reduced mean absolute age error from 0.68 ± 0.01 to 0.52 ± 0.01 years and increased sex accuracy from 81.3 ± 1.7% to 85.8 ± 0.8%. Adding 20 deciduous-tooth classes provided no age benefit and reduced sex accuracy by 7.9% relative to the permanent-only representation. The mandibular first molars received the greatest tooth-level attention, and their occlusion impaired both prediction tasks. These findings show that anatomically plausible inputs do not necessarily provide incremental predictive value. Task-specific input auditing can improve joint model performance, reduce input redundancy and preserve anatomically traceable predictions. More broadly, this framework provides a testable approach for selecting correlated inputs in medical-imaging systems in which regional views re-present anatomy already contained in the whole image.

## Introduction

Estimating chronological age and sex from dental radiographs supports forensic identification and legal assessment when reliable documents are unavailable^1–4^. Age thresholds are particularly consequential: in China, Criminal Law Amendment XI extends criminal responsibility to individuals aged 12–14 years for narrowly specified serious offences, subject to approval by the Supreme People’s Procuratorate^5^. Near such boundaries, calibrated threshold probabilities are more informative than average estimation error alone^6,7^. A single orthopantomogram (OPG) captures the full dentition and surrounding maxillofacial structures^8^. Dental development follows a relatively ordered, although individually variable, sequence^9–13^, making teeth informative for age estimation in children and adolescents. The same image also contains dental and craniofacial morphology relevant to sex estimation^14–18^. An OPG therefore provides a common anatomical basis for joint age and sex modelling.

Conventional dental age assessment relies on stage-based systems, including the Demirjian, Willems, Nolla and Cameriere methods^9,19–22^. These approaches are well established but require expert tooth staging and remain sensitive to observer variation^23,24^. Deep learning has shifted the task toward automated feature extraction from OPGs. Such networks now reach clinical-grade performance in several imaging tasks^25^, and graph-based representations support diagnosis and report generation^26,27^. Whole-image models estimate age directly from panoramic radiographs, sometimes using sex as an auxiliary input^28–36^, whereas tooth-centred approaches classify developmental stages or analyse detected tooth regions^37–41^. Multi-task models further estimate age and sex from the same radiograph^15^. Together, these approaches move beyond manual scoring, but they differ substantially in the anatomy presented to the network. Input selection therefore becomes a central methodological choice: whole images preserve global craniofacial context, whereas cropped jaws and individual teeth provide progressively more local representations of anatomy already contained in the same exposure.

Multi-input learning is widely used in medical imaging to combine imaging sequences, complementary modalities, clinical data or anatomically defined regions^42–48^. Its benefit is generally attributed to the complementary information supplied by each input. This premise becomes less secure when candidate inputs are derived from a single image and present the same anatomy at different scales. Multimodal networks can underperform their strongest single-input counterpart^49^, and theoretical and empirical studies show that added views are useful only under specific relationships with the information already retained^50–55^. In OPG analysis, the panoramic image, a jaw-region crop and individual tooth regions are spatially nested rather than independent views. Their fused performance therefore cannot establish the incremental value of each input or show whether its utility differs between age estimation and sex classification^44,56,57^. Existing OPG models demonstrate the feasibility of whole-image, tooth-region and multi-task approaches^15,28,37^, but do not isolate the task-specific contribution of overlapping anatomical inputs. The unresolved question is therefore not how much anatomy can be supplied to a model, but which inputs provide incremental information and which should be removed as redundant.

In this work, we develop HMA-Net together with a task-specific input audit to determine whether anatomically nested representations provide incremental utility for joint age estimation and sex classification from a single orthopantomogram (Fig. 1a). The framework evaluates the panoramic image, jaw-region crop and detected tooth regions as candidate inputs, retaining a stream only when it improves at least one task without materially degrading the other. HMA-Net represents individual teeth as an anatomically indexed graph and uses task-specific feature-wise linear modulation to preserve the separability of the input streams and prediction tasks. Because attention weights alone do not establish functional relevance^58–60^, we complement tooth-level attention with cross-stream occlusion analyses^61–63^. The audit identifies the panoramic image and permanent- tooth regions as the retained input set: the jaw-region stream degrades both outcomes once the tooth stream is included, whereas deciduous-tooth regions provide no age benefit and reduce sex-classification performance. Attention and occlusion analyses further identify the mandibular first molars as regions relevant to both predictions. These findings show that task-specific input auditing can produce a more parsimonious and anatomically traceable model and may inform medical-imaging systems in which candidate inputs represent overlapping anatomy at different scales.

**Fig. 1:**
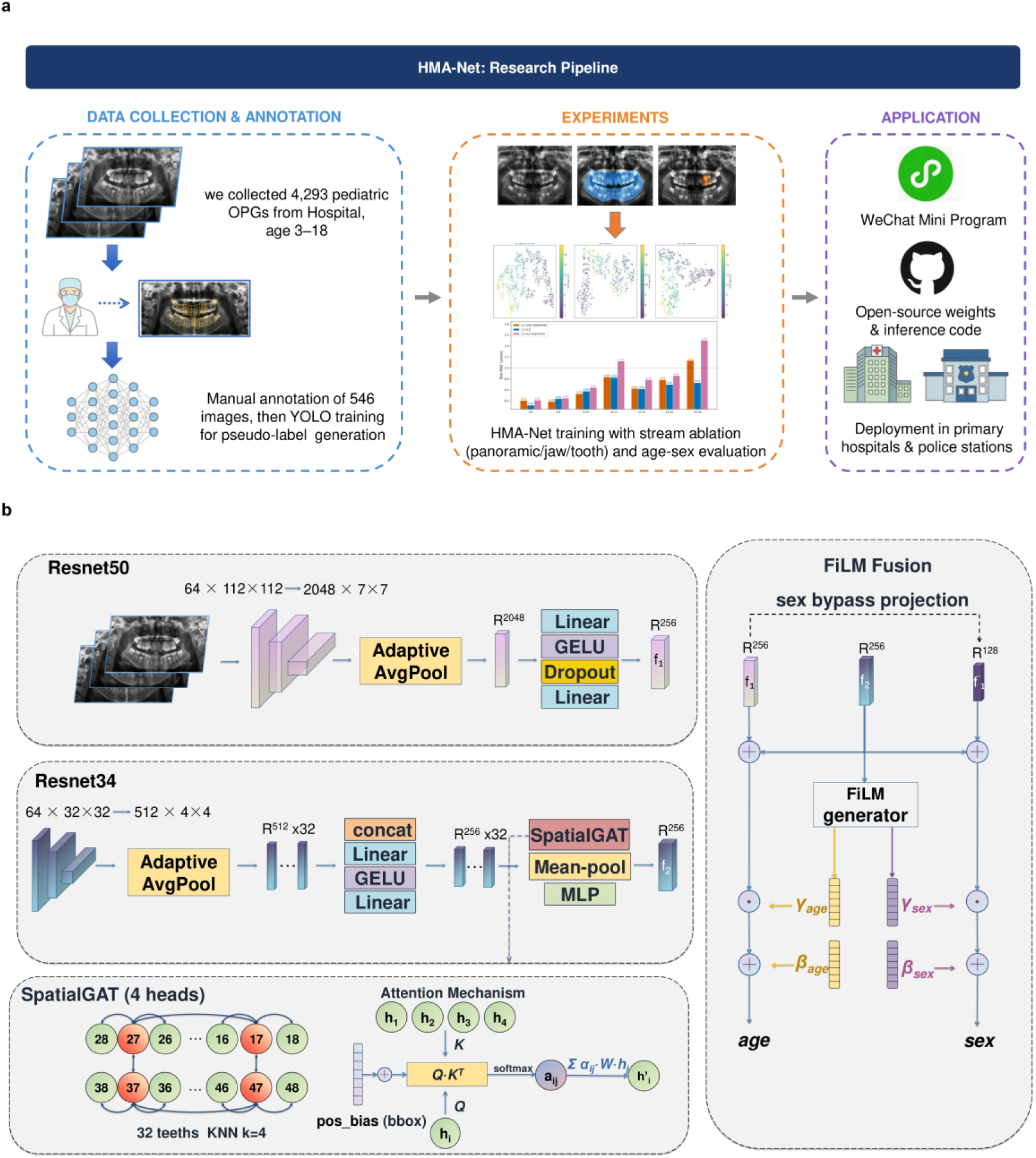
HMA-Net overview. (a)Research pipeline, from data collection to the intended forensic and clinical application. (b) Model architecture: a panoramic stream (L1, ResNet50) and a per-tooth graph stream (L3, ResNet34 with a k-NN graph attention network) are fused by FiLM modulation into co-equal age and sex heads.

## Results

### HMA-Net integrates global and tooth-level information

We developed HMA-Net as an anatomy-aware two-stream architecture for joint chronological age estimation and sex classification from a single orthopantomogram (OPG; Fig. 1). A ResNet^60^ panoramic stream encoded global craniofacial information, whereas a shared ResNet34 extracted local features from 32 permanent-tooth regions of interest. Each tooth representation was linked to its FDI identity and integrated with neighbouring teeth using a four-neighbour graph attention network. Task-specific feature-wise linear modulation then fused the panoramic and tooth representations into separate age-regression and sex-classification heads. The dataset comprised 4,293 OPGs from participants aged 3–18 years and was partitioned by subject into training (n = 2,999), validation (n = 637) and test (n = 657) sets (Fig. 2a).

**Fig. 2:**
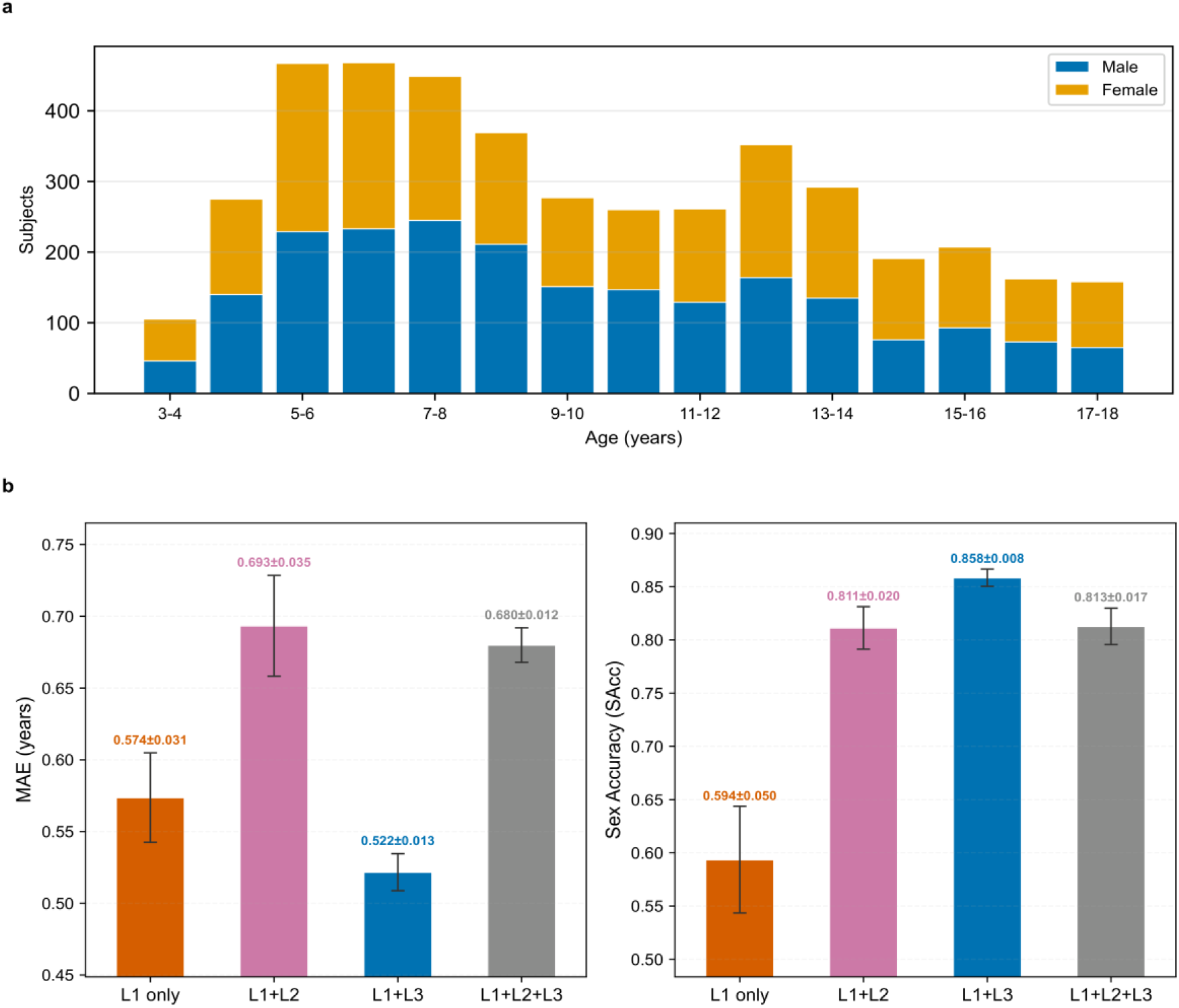
Study cohort and stream configuration. (a) Age–sex distribution of the 4,293-subject cohort. (b) Age MAE and sex accuracy (mean ± SD, three runs) for four stream configurations; L1+L3 is the best-balanced configuration.

On the held-out test set, the final panoramic-plus-permanent-tooth model achieved an age mean absolute error (MAE) of 0.33 years and a sex-classification accuracy of 85.2% (Supplementary Table 1). Overall, 99.4% of the estimated ages were within 2 years of chronological age. Unless otherwise stated, configuration comparisons below are reported as the mean ± standard deviation across three independent training runs, whereas the detailed error, threshold and occlusion analyses were performed using the final model.

### Removing redundant inputs improves joint-task performance

We first evaluated the incremental utility of the three candidate anatomical inputs: the full panoramic image (L1), a jaw-region crop (L2) and individually detected permanent-tooth regions (L3). Under identical training and evaluation settings, the panoramic-only model achieved an age MAE of 0.574 ± 0.031 years and a sex-classification accuracy of 59.4% ± 5.0% (Fig. 2b). Adding the permanent-tooth stream reduced the MAE to 0.522 ± 0.013 years, corresponding to a relative reduction of 9.1%, and increased sex accuracy to 85.8% ± 0.8%, an absolute increase of 26.4 percentage points. Both differences were supported by pooled comparisons across the three runs (two-sided Mann–Whitney U test for absolute age errors, P < 0.001; Fisher’s exact test for sex classification, P < 0.001). The permanent-tooth stream was therefore retained because it improved both prediction tasks.

The jaw-region stream did not provide the same joint benefit. The panoramic-plus-jaw model achieved an age MAE of 0.693 ± 0.035 years and a sex accuracy of 81.1% ± 2.0%. Although the jaw crop improved sex classification relative to the panoramic-only model, it increased age MAE by 20.7%. Moreover, adding the jaw crop to the panoramic-plus-tooth model increased age MAE from 0.522 ± 0.013 to 0.680 ± 0.012 years and reduced sex accuracy from 85.8% ± 0.8% to 81.3% ± 1.7%. The corresponding increases in age error and reductions in sex accuracy were supported by the pooled comparisons (both P < 0.001). The jaw-region stream was therefore excluded because it provided no complementary benefit once the permanent-tooth stream was present.

We next examined whether panoramic-image resolution influenced the contribution of the retained streams. Increasing the panoramic resolution from 256 × 256 to 512 × 512 pixels reduced the age MAE of the panoramic-plus-tooth model from 0.622 to 0.522 years and increased sex accuracy from 80.5% to 85.8% (Supplementary Fig. 1). In the panoramic-only model, the same increase in resolution reduced age MAE from 0.657 to 0.574 years but decreased sex accuracy from 62.2% to 59.4%. The 512 × 512 resolution was therefore retained for the final two-stream model.

Finally, we compared a tooth stream comprising 32 permanent-tooth classes with an expanded representation that additionally included 20 deciduous-tooth classes. Adding the deciduous-tooth classes produced no detectable difference in age estimation across the full test set (mean difference, 0.002 years; 95% CI, −0.2 to 0.02; paired Wilcoxon signed-rank test, P = 0.60) but reduced sex accuracy by 6.5 percentage points (95% CI, −10.0 to −3.2; exact McNemar test, P < 0.001). Accordingly, the retained input configuration comprised the 512 × 512 panoramic image and 32 permanent-tooth regions, without the jaw-region crop or deciduous-tooth classes.

### Component selection trades age error against sex accuracy

With the panoramic and permanent-tooth streams fixed, we compared alternative fusion, aggregation, attention, and backbone components at the same age-model checkpoint (Supplementary Fig. 2). Feature-wise linear modulation (FiLM) produced a lower age MAE and slightly higher sex accuracy than the gating baseline. Cross-attention and multimodal transfer module fusion lowered age error below the gating baseline (0.639 and 0.606 vs 0.643 years) but reduced sex accuracy (61.3% and 69.1% vs 75.7%), whereas replacing graph attention with average pooling worsened both outcomes. FiLM fusion with graph attention therefore provided the most favourable balance between the two tasks among the evaluated component variants.

Graph connectivity produced a similar trade-off (Supplementary Fig. 3). With four neighbours per tooth, the model achieved an age MAE of 0.622 years and a sex accuracy of 76.3%. Reducing the neighbourhood to two neighbours increased MAE to 0.647 years and slightly reduced sex accuracy to 75.7%. Increasing the neighbourhood to six or eight neighbours produced MAEs of 0.641 and 0.644 years and sex accuracies of 68.7% and 73.4%, respectively. A fully connected graph achieved the lowest MAE in this analysis, 0.607 years, but reduced sex accuracy to 66.4%. The four-neighbour graph was retained because it avoided the larger loss in sex performance observed with denser graphs while maintaining a lower age error than the other k-nearest-neighbour settings.

The loss-function analysis showed why both outcomes had to be considered together (Supplementary Fig. 4). Training with mean squared error alone reduced age MAE from 0.622 to 0.605 years but left sex accuracy near chance at 50.4%. Combining mean squared error with standard binary cross-entropy increased sex accuracy to 77.9% but also increased age MAE to 0.654 years. Focal binary cross-entropy yielded an age MAE of 0.622 years and a sex accuracy of 76.3%. It was retained because it preserved both tasks without the larger deterioration observed with either alternative.

Among the seven evaluated augmentation schedules^64^, the fixed-mid strategy occupied the most favourable joint position, with the lowest plotted age MAE and the highest sex accuracy (Supplementary Fig. 5). Descriptive t-SNE visualisations also showed clearer age ordering and sex separation in the fused panoramic-plus-tooth representation than in either single-stream representation (Supplementary Fig. 6). These analyses defined the retained model as the 512 × 512 panoramic-plus-permanent-tooth configuration with FiLM fusion, four-neighbour graph attention, focal binary cross-entropy, and fixed-mid augmentation. Each component was retained on the same joint basis applied throughout this section, a gain for at least one task with no task falling beyond the seed-to-seed variation; the input-utility form of this criterion is formalised in the Methods.

### Age errors increase and threshold errors cluster near decision boundaries

Age-estimation error increased across the seven age groups in all four input configurations (Fig. 3a). The panoramic-plus-tooth model had the lowest MAE in each group. At 12–14 years, its MAE was 0.56 years, compared with 0.56 years for the panoramic-only model; in the 16–18-year group, the corresponding values were 1.03 and 1.08 years. Configurations containing the jaw-region stream showed higher age errors throughout the age range, with the largest differences in the oldest group. The seven groups contained 129, 140, 99, 80, 99, 61, and 49 participants, respectively, leaving fewer observations in the two oldest groups.

**Fig. 3:**
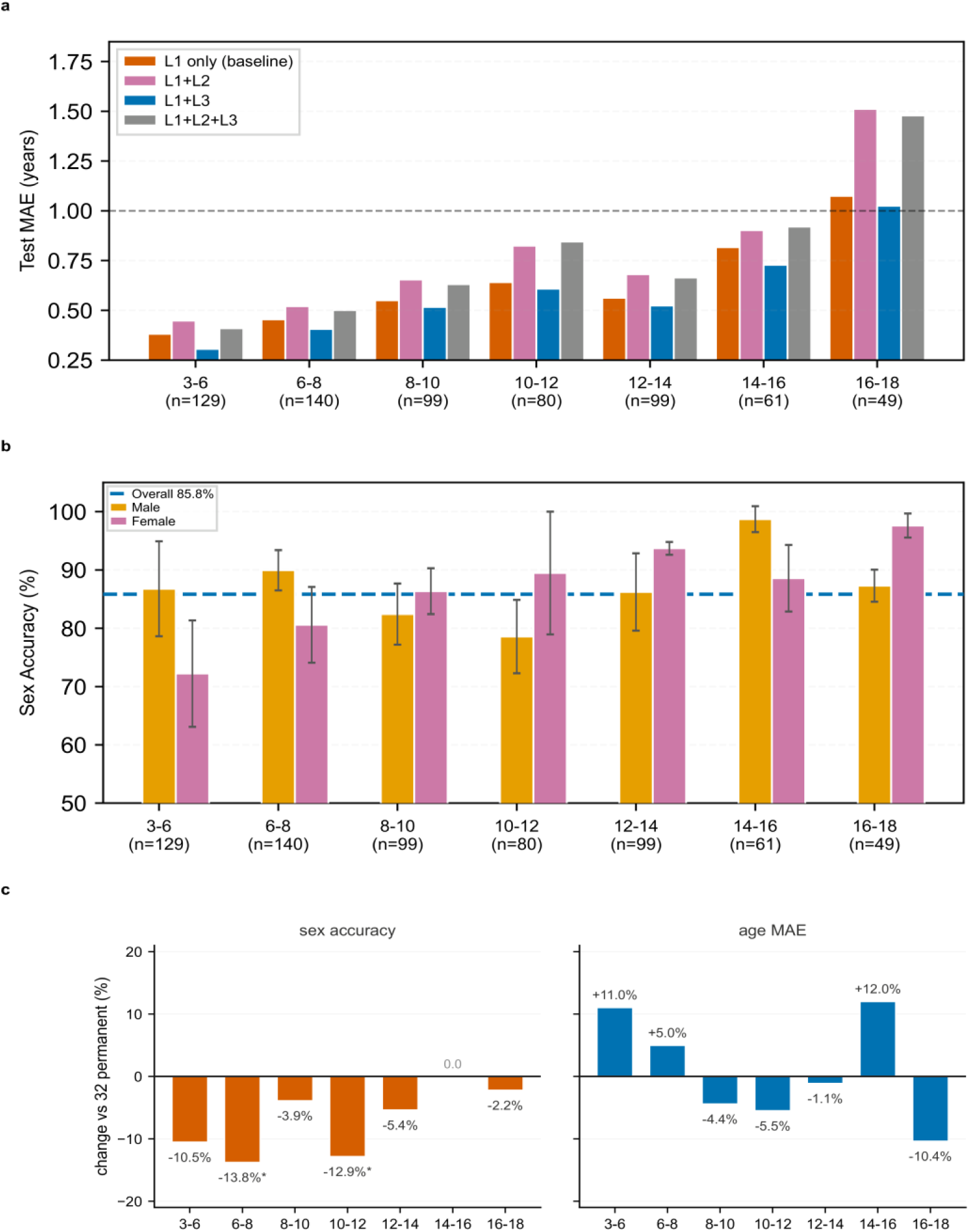
Input-source and per-age-bin analysis. (a) Per-age-bin age MAE and (b) per-age-bin sex accuracy for the four input configurations; n = 49–140 subjects per bin and means ± SD across three runs. (c) Relative change from expanding the tooth stream to 52 classes (20 deciduous classes added, 32-permanent configuration as reference), computed on the same subjects of each bin and plotted on a common percentage scale; asterisks mark bins with a significant paired McNemar test on sex accuracy.

Sex-classification accuracy also varied across age groups and between sexes (Fig. 3b). Accuracy was lowest in the 3–6-year group (79.3%) and higher beyond 10 years, although the pattern was not monotonic. Female accuracy was higher in the 8–10, 10–12, 12–14 and 16–18-year groups, whereas male accuracy was higher in the 3–6, 6–8 and 14–16-year groups. Neither sex therefore showed consistently higher classification accuracy across all developmental stages.

Across the full test set, absolute age errors were concentrated below 1 year, and 99.4% of predictions were within 2.0 years of chronological age (Fig. 4a). Predicted age followed chronological age closely across the evaluated range, although the dispersion increased among older participants (Fig. 4b). We therefore examined whether this overall agreement translated into reliable decisions at the prespecified age thresholds.

**Fig. 4:**
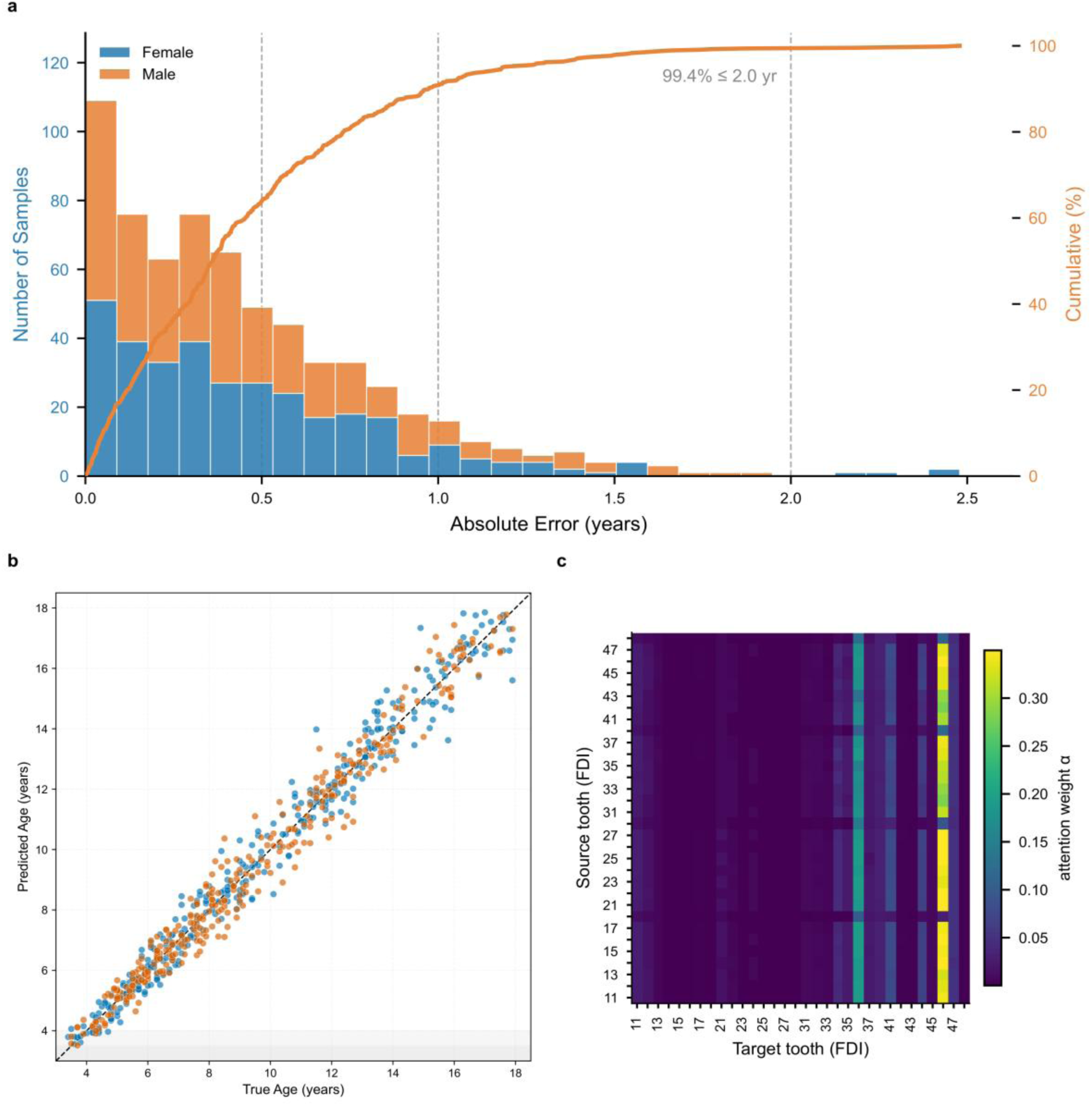
Error characterisation. (a) Error histogram with cumulative distribution. (b) Predicted versus true age, coloured by sex. (c) GAT edge-attention matrix (32 × 32) indexed by FDI tooth number.

At the 12-, 14-, and 16-year thresholds, the areas under the receiver operating characteristic curves were 0.993, 0.994, and 0.995, respectively (Fig. 5a). In the binned calibration analysis, predicted ages remained close to the corresponding observed mean ages. Observed-minus-predicted differences ranged from +0.11 years in the youngest prediction bin to −0.17 years in the oldest bin (Fig. 5b). Median prediction bias remained close to zero across the age groups, although the dispersion of individual errors was larger in the older groups (Fig. 5c).

**Fig. 5:**
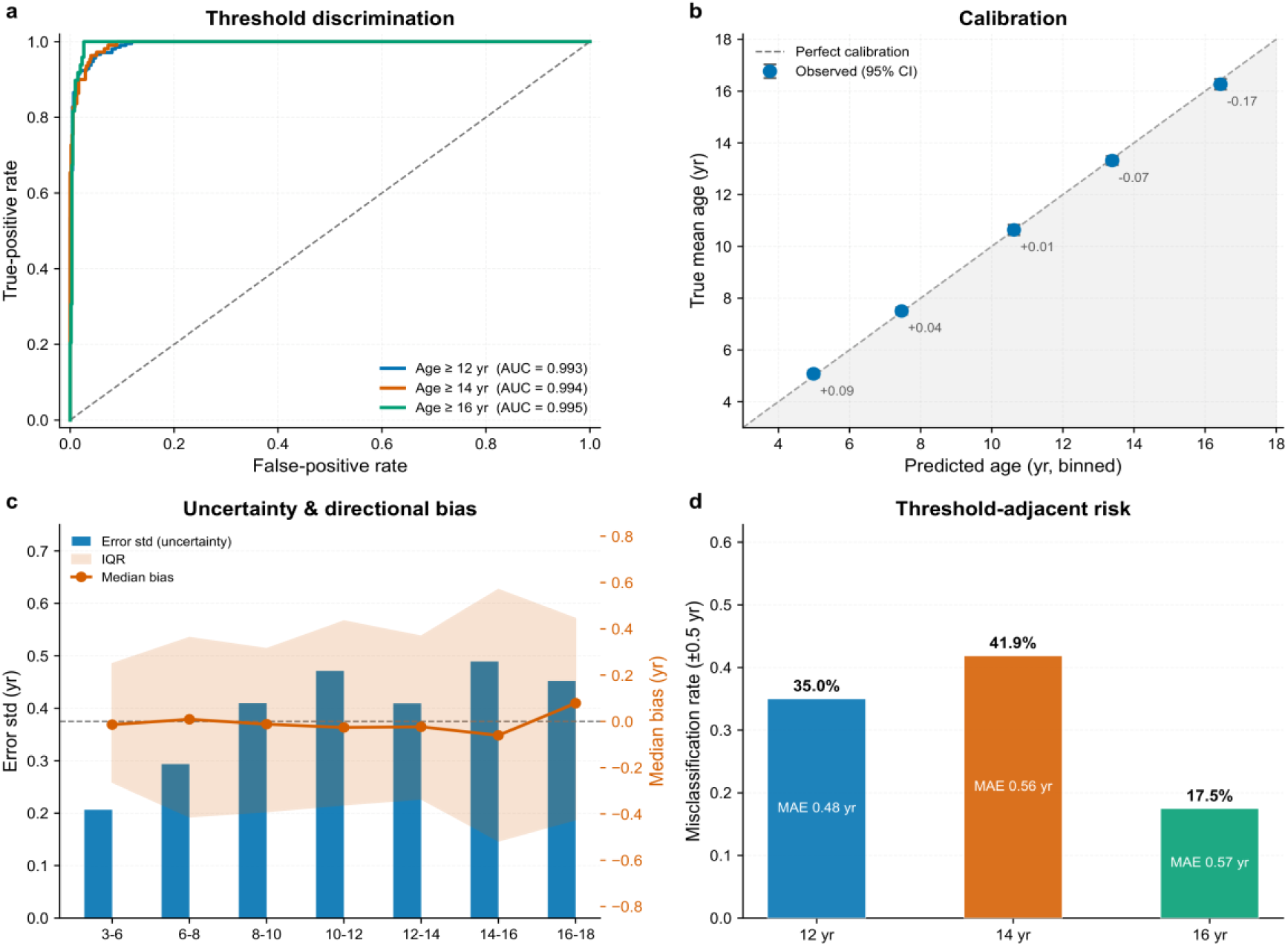
Forensic threshold analysis. (a) ROC curves for the age ≥ 12, 14 and 16 yr thresholds. (b) Calibration: binned predicted versus true mean age. (c) Uncertainty and directional bias by age bin. (d) Misclassification rate within ± 0.5 yr of each legal threshold.

Performance was less consistent close to the decision boundaries. Within ± 0.5 years of the 12-, 14-, and 16-year thresholds, the misclassification rates were 35.0%, 41.9%, and 17.5%, respectively (Fig. 5d). The corresponding age MAEs within these boundary regions were 0.48, 0.56, and 0.57 years. The highest local misclassification rate occurred around the 14-year threshold despite the high AUC observed across the full test set.

Descriptive threshold estimates also varied by sex. At 12 years, the AUC was 0.994 for female participants and 0.992 for male participants; the corresponding values were 0.989 and 0.997 at 14 years and 0.995 and 0.994 at 16 years (Supplementary Table 2). Within ± 0.5 years of each threshold, female and male misclassification rates were 32.1% and 37.5% at 12 years, 63.6% and 19.0% at 14 years, and 22.2% and 13.6% at 16 years. The largest observed separation was concentrated around the 14-year boundary. These findings show that high overall discrimination did not translate into equally low error among individuals whose chronological ages were close to a decision threshold.

### Mandibular first-molar attention is supported by occlusion sensitivity

The final analysis asked whether tooth-level attribution identified regions to which the model was functionally sensitive. The graph representation assigned attention to individual teeth by FDI number rather than only to image regions. The mandibular first molars received the largest aggregate incoming attention: FDI 46 accounted for 0.51 and FDI 36 for 0.17, both exceeding the values assigned to the remaining teeth (Fig. 6a). The 32 × 32 edge-attention matrix similarly showed prominent incoming-attention bands for these two target teeth (Fig. 4c). Grad-CAM maps provided complementary spatial localisation in the panoramic and tooth inputs, whereas the graph representation retained the identity of each attributed tooth (Fig. 6b).

**Fig. 6:**
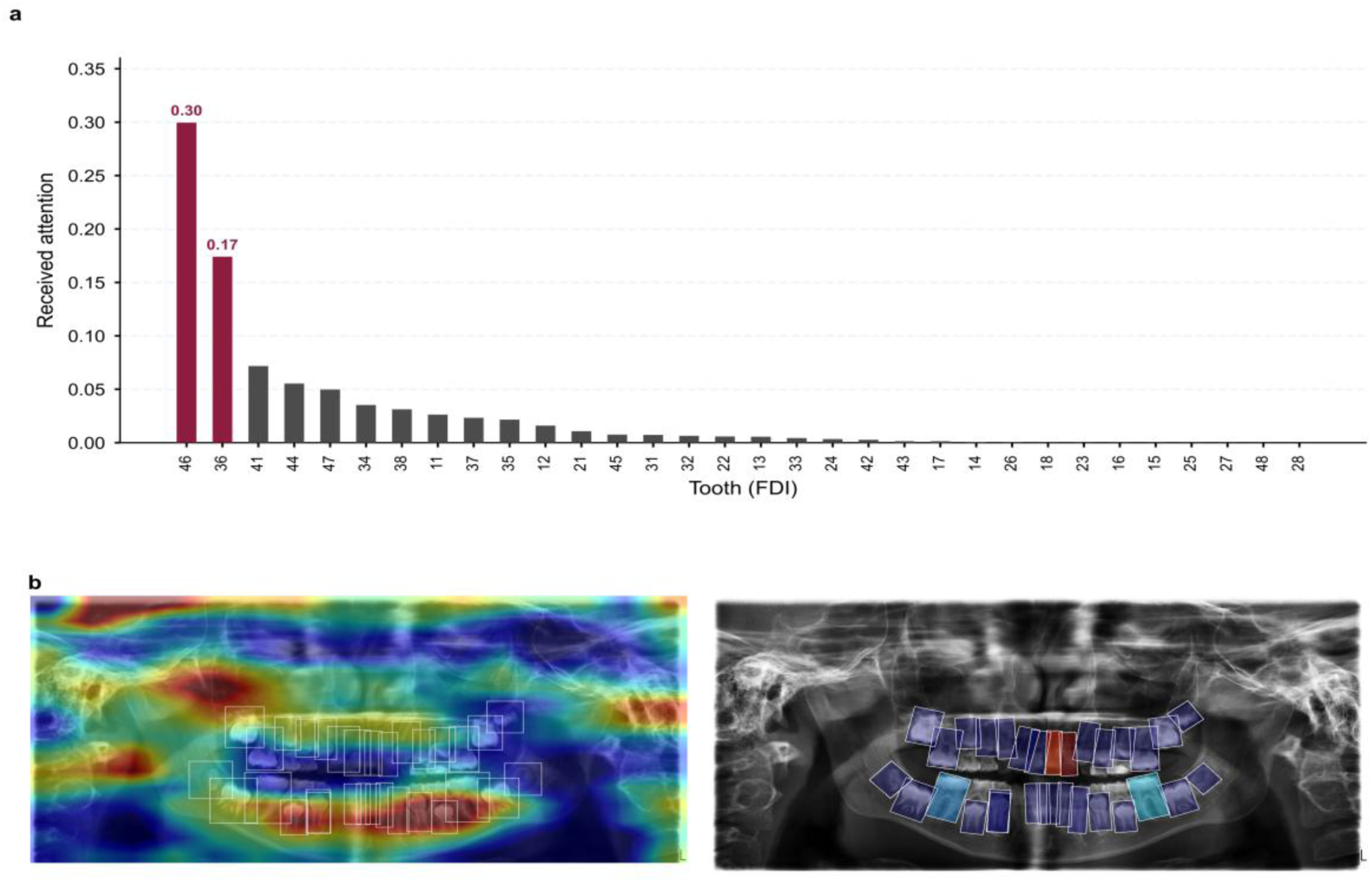
Attention-based interpretability. (a) Per-tooth received GAT attention, sorted by magnitude. (b) Grad-CAM attribution for the panoramic and per-tooth streams.

The two highest-attention teeth were occluded in both the panoramic and tooth streams. With a square mask whose side length was 1.4 times that of the detected tooth box, simultaneous occlusion of FDI 36 and 46 increased age MAE from 0.505 to 0.694 years and root mean squared error from 0.668 to 0.869 years, while sex accuracy decreased from 84.6% to 81.6% (Supplementary Fig. 7); the occlusion harness performs its own forward pass, so its unoccluded reference (MAE 0.505 years, sex accuracy 84.6%) differs marginally from the main evaluation pipeline (0.51 years, 85.2%). Occluding FDI 36 alone increased MAE to 0.558 years and reduced sex accuracy to 82.6%; occluding FDI 46 alone increased MAE to 0.563 years and reduced sex accuracy to 83.7%. The larger degradation after masking both molars was consistent with the concentration of received attention on these teeth.

The increase in age error became larger as the mask was enlarged, while sex accuracy declined at every mask size. When both molars were occluded with masks ranging from 1.0 to 2.0 times the tooth-box side length, the increases in MAE were 0.087, 0.158, 0.189, 0.257, and 0.562 years, respectively; the corresponding reductions in sex accuracy were 1.8, 4.0, 3.0, 4.9, and 10.5 percentage points (Supplementary Fig. 7a). In the complementary mask-geometry analysis, occluding both molars increased MAE by 0.066 years with a tight mask, 0.189 years with a 1.4-fold mask, and 0.818 years with a 2.24-fold mask (Supplementary Fig. 7b). Masking the mandibular first molars degraded both tasks, indicating that the predictions rely on these regions. The attention weights, however, may not explain how the model reaches its decision: the masks were square and grew with the tooth box, so larger masks also removed neighbouring teeth and bone, and part of the performance drop may therefore reflect collateral damage to neighbouring structures in addition to the molars themselves.

## Discussion

This study used a structured input audit to determine which anatomical representations contributed to joint age estimation and sex classification from paediatric orthopantomograms. The smaller panoramic-plus-permanent-tooth model provided the best joint-task balance: the permanent-tooth stream improved both outcomes, whereas the jaw-region stream and deciduous-tooth classes did not add consistent complementary value. Once the input set was fixed, FiLM fusion and graph attention maintained a more favourable balance between the two tasks than the evaluated alternatives. Performance nevertheless remained heterogeneous across development and near the prespecified age thresholds, and the tooth-level attribution analyses identified the mandibular first molars as regions to which the model was particularly sensitive under occlusion. These findings support the value of evaluating each candidate input against the streams already retained rather than assuming that additional anatomically defined inputs will improve a multi-input model.

The input ablations suggest that anatomical plausibility alone does not guarantee incremental predictive utility. The jaw-region crop improved sex classification relative to the panoramic-only model but increased age error, and adding it to the panoramic-plus-tooth configuration degraded both outcomes. One possible explanation is that the crop re-presented information already available in the panoramic image without adding the explicit geometric measurements used in conventional mandibular assessment, such as ramus height or gonial angle^16,65^. The deciduous- tooth classes may have introduced a different form of redundancy by contributing less consistent signals across the 3–18-year age range. These explanations remain hypotheses because the present experiments measured predictive performance rather than the information content of each stream. By contrast, the permanent-tooth stream preserved local crown and root-development features that were represented at lower resolution in the whole-image stream. More broadly, this pattern is consistent with evidence that correlated inputs can compete during joint training and that adding another view does not necessarily improve a multimodal model^50,51, 49,52^.

The architecture ablations further showed why the two outcomes had to be evaluated together. FiLM fusion with graph attention was associated with the most favourable joint-task performance among the tested component variants, whereas four of the 17 component substitutions (cross-attention, MMTM, ConvNeXt-T and the ResNet^66^ panoramic backbone) reduced age error at the cost of sex accuracy. The same trade-off appeared in the topology and loss analyses: a fully connected graph and mean-squared-error-only training achieved lower age errors in their respective comparisons but substantially weakened sex classification. The four-neighbour graph was therefore selected for its joint performance rather than because it was uniformly optimal for either task. Its two-hop receptive field covering approximately one dental quadrant provides an anatomical rationale for this setting. Similarly, FiLM may help preserve task-specific information and graph attention may help integrate relations between teeth.

Performance across age groups and decision thresholds defines an important boundary of the model. Age-estimation error increased in the older groups, where dental maturation is closer to completion and individual developmental variation may become more influential^9–13;^ the smaller sample sizes in the two oldest groups may also have contributed to less stable estimates. Although discrimination at the 12-, 14-, and 16-year thresholds was high across the full test set, errors were concentrated among participants whose chronological ages lay close to the decision boundaries. Global AUC therefore did not imply equally reliable classification for the individuals in whom the threshold decision was most difficult, consistent with the distinction between overall regression performance and threshold-specific assessment^6,7,67^. The largest descriptive difference between female and male participants occurred around the 14-year threshold, but this subgroup result should not be interpreted as a stable sex effect without confidence intervals and formal interaction testing. For forensic use, threshold-specific error, uncertainty, and calibration should therefore accompany the overall MAE and AUC rather than being inferred from them.

The FDI-indexed graph provided a complementary form of interpretability by assigning attention to named teeth and tooth-to-tooth relations rather than only to image regions. The mandibular first molars received the highest aggregate incoming attention, which is compatible with the established importance of molar development in dental age assessment^10,12,19^. Occluding these teeth in both input streams degraded age and sex performance, and simultaneous occlusion produced a larger deterioration than single-tooth occlusion. This agreement between attention ranking and perturbation response supports functional sensitivity to the mandibular first-molar regions. It does not establish that the attention weights are a causal explanation^58,61^. Black-mask occlusion creates an artificial input, and larger masks also modify information surrounding the target tooth. Future analyses should include area-matched random-tooth and non-dental controls, texture-preserving or counterfactual perturbations, and stability analyses across training runs and external cohorts.

The final model’s age error was within the range reported for manual staging and previous deep-learning studies in paediatric cohorts^3^, 14, 15, 58-60, 70 (Supplementary Table 3), but differences in age distribution, image acquisition, reference labels, data splitting, and evaluation protocols preclude a direct ranking of methods. The main contribution is therefore not a claim of universal performance superiority, nor a new mathematical theory of input value, but a structured procedure for testing the incremental contribution of anatomically overlapping inputs for each task. This question may also be relevant to other medical-imaging settings in which a region of interest is nested within a larger image or when inputs re-present correlated anatomy. Whether the same audit remains informative in CT, MRI, or other multimodal applications requires independent evaluation; the present findings establish its use only for the OPG configurations and outcomes examined here.

The strengths of this study are a paediatric cohort spanning primary eruption to late adolescence, age and sex labels for every participant, controlled comparisons of overlapping anatomical inputs, and attribution analyses that retained tooth identity. Five limitations constrain the conclusions. The data were obtained from a single centre and a Chinese Han population, with greater representation of younger children, and external multicentre validation is required. The oldest age groups and the sex-stratified threshold regions contained relatively few participants, and the absence of confidence intervals and interaction tests limits subgroup inference. The evaluated age range ended below 18 years, leaving the 18-year threshold and adult performance unassessed. The tooth stream also depended on detector-generated boxes and pseudo-labels, so detection errors could propagate into both prediction and attribution. The input audit estimated first-order marginal effects for specific parent configurations and did not capture all higher-order or order-dependent interactions among streams. Future work should evaluate a locked model in an independent external cohort not used in model development^68,69^, quantify uncertainty for the threshold and occlusion analyses, and test the audit prospectively in other populations and imaging settings.

In this study, a structured input audit was used to determine whether anatomically overlapping representations added task-specific value to joint age estimation and sex classification from paediatric orthopantomograms. The retained panoramic-plus-permanent-tooth configuration achieved a more favourable joint-task balance than configurations containing a jaw-region stream or deciduous-tooth classes, showing that removing inputs without consistent incremental value can improve input efficiency and, in this dataset, predictive performance. Tooth-level attribution and occlusion experiments further indicated that the model was particularly sensitive to the mandibular first-molar regions. However, age error increased in older participants, and misclassification rates were relatively higher near the evaluated age thresholds despite high overall discrimination. These findings support systematic evaluation of candidate inputs rather than their automatic inclusion. More broadly, this input-audit perspective may inform the development of medical imaging diagnostic and prediction models in which multiple inputs represent overlapping anatomy. Validation of a locked model in independent populations, together with further assessment of threshold-specific uncertainty, is required before forensic application.

## Methods

### Data and Study Cohort

Study cohort. Joint age-and-sex estimation requires a dataset spanning the full paediatric developmental window with both labels for every subject, which existing public OPG datasets do not provide: they are predominantly adult-focused or lack sex labels. We therefore assembled 4,293 orthopantomograms (OPGs) of paediatric patients aged 3–18 years at Nanfang Hospital, Southern Medical University, covering primary eruption through permanent completion. Each image carries chronological age and sex labels; the age distribution peaks at 5–7 years and sex is balanced (2,156 female, 2,137 male). The dataset was split by subject into training, validation and test sets of 2,999, 637 and 657 subjects (Fig. 2a). All results are reported on the 657-subject held-out test set. The study was retrospective and used a fixed subject-level partition: no randomization was applied when assigning subjects to the training, validation and test sets, and no blinding was used, because chronological age and sex were extracted from the clinical record before any model development. Sex was recorded as a binary clinical variable for every subject and served as the classification target; gender identity was not recorded in the source data.

Preprocessing. Each OPG was resized to 512 × 512 for the panoramic stream, and the 32 permanent-tooth regions were cropped at 64 × 64 after per-tooth detection; pixel intensities were normalised to [0, 1]. Positions without a detected tooth were zero-filled and marked invalid by a binary mask; they were excluded from graph attention and from the mean pooling that forms the tooth-stream feature.

### Tooth Detection Module

#### Detector selection

We evaluated eight YOLO OBB variants (YOLO8n through YOLO11l)^70^ on a manually annotated test set (Supplementary Fig. 8). YOLO8m-OBB gave the highest mAP@50 (0.977) and was selected for its balance of accuracy and inference speed. Detection used the Ultralytics implementation (8.4.10), 1024-px input, a confidence threshold of 0.25, NMS IoU of 0.7 and at most 300 boxes, keeping the highest-confidence box when more than one was returned for the same FDI position. Detector variants were trained for up to 300 epochs at 1024-px input with cosine decay and early stopping (patience 15; batch size 4 for the medium variant); full configurations accompany the code repository.

#### Tooth annotation and pseudo-labels

The detector distinguishes 52 tooth classes (32 permanent, 20 deciduous) and supplies the 32 per-tooth ROIs that feed the tooth stream. We manually annotated 546 images with 52-class oriented bounding boxes in FDI numbering, labelling a tooth whenever its structure was visible, erupted or not; absent teeth were not annotated, and restored teeth were annotated as any other tooth. A YOLO8m-OBB detector trained on those images generated pseudo-labels for the remaining 3,747. HMA-Net consumes only the 32 permanent classes: the 20 deciduous classes are transient within a 3–18 year window and did not improve age MAE while degrading sex accuracy (Results). Detector-predicted boxes were used for both training and inference, so the model sees realistic detector noise; the label pool holds 4,299 OPGs, six more than the analysed cohort, and the graph-topology analysis uses that full pool (Supplementary Table 4).

### Model Architecture

#### Model specification

HMA-Net is a supervised two-stream network that we constructed for joint age regression and sex classification from OPG images. Fusion modules and prediction heads are trained from scratch; the ResNet^60^ and ResNet34 backbones are ImageNet-initialised. Inputs are a 512 × 512 greyscale OPG and 32 × 64 × 64 greyscale tooth ROIs; outputs are a continuous age estimate (years) and a binary sex class. The model targets paediatric forensic and orthodontic use; performance on adults has not been established.

#### Two spatial scales

HMA-Net analyses the OPG at the two scales at which age and sex are signalled, fused by FiLM (Fig. 1): L1 for global craniofacial context and L3 for local developmental features. Craniofacial proportions (jaw shape, cranial width) carry sex-discriminative and coarse age information at the full-image scale^71^, whereas the cues dental staging relies on, root apex closure and crown morphology, exist at the level of individual teeth and are destroyed by the downsampling needed for the whole image. The effective receptive field of a CNN is much smaller than its theoretical one^72,73^. Encoding each scale separately therefore preserves information that a single backbone loses, and anatomical decomposition injects domain knowledge that helps on a cohort of 4,293 subjects^74^. Each candidate input is treated as a hypothesis rather than a default: a jaw-crop stream was tested on that basis and discarded (Results).

The panoramic stream (L1) encodes global craniofacial context at 512 × 512 with a ResNet50 backbone^71,75^, giving a 256-dimensional vector. The deeper encoder is warranted because arch shape and sinus morphology are global features, and this stream carries most of the sex signal.

The tooth stream (L3) processes 32 tooth ROIs at 64 × 64 through a shared ResNet34 backbone^71^, preserving detail that the panoramic encoding cannot resolve (Results). To tie each feature to an anatomical identity rather than to image content alone, the 512-dimensional backbone output is concatenated with a 32-dimensional embedding of the ROI’s oriented bounding box and a 16-dimensional FDI embedding, and projected to 256 dimensions. This inductive bias fixes every feature to an explicit FDI index, which is what makes tooth-level attribution possible (Results).

The 32 per-tooth features form a sparse graph in which each tooth is connected to its k = 4 spatial nearest neighbours, computed from tooth-centre coordinates. This KNN topology, chosen by systematic ablation (Results) and by a receptive-field analysis on the detected boxes (Supplementary Table 4), encodes the expectation that anatomically adjacent and opposing teeth supply the most relevant developmental context. A four-head, two-layer graph attention network (GAT)^66^ processes the graph and produces updated node representations (Eq. 1):

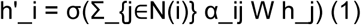

where α_ij are learned attention coefficients and W a learnable weight matrix. The attention logits add a learned bias from each tooth’s bounding-box coordinates, so the KNN mask decides which teeth may interact while the position bias modulates how strongly each interaction is weighted, following the arch geometry. The updated nodes are mean-pooled into the 256-dimensional tooth-stream feature f_3.

#### Denoising adapter

Because the ROIs come from an automated detector, the pooled tooth feature carries detection-level noise. A residual MLP with layer normalisation (256 → 256 → 256; linear, layer norm, GELU, dropout, linear) maps it toward a cleaner latent space before fusion, preserving the 256-dimensional size.

### FiLM Fusion and Multi-Task Heads

Given f_1 ∈ R^256 (L1) and f_3 ∈ R^256 (L3), the two streams are fused by FiLM (Feature-wise Linear Modulation)^76^. Unlike concatenation or addition, which treat the streams as independent contributors, FiLM applies a learned channel-wise affine transform to their concatenation, letting tooth-level developmental features condition how the fused representation is modulated (Eqs. 2 and 3):

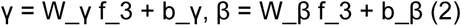

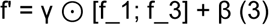

The modulated representation is routed to two heads: a single linear layer trained with MSE for age, an ordinal-aware variant^77^ not being adopted because of its worse calibration, and a sigmoid-activated linear layer trained with focal BCE^78^ (γ = 2) for sex. Focal loss emphasises hard examples under class imbalance.

Fusion is task-decoupled, treating age and sex as co-equal targets^79^. Two independent FiLM generators, one per task, each produce channel-wise scale and shift from f_3: the age path modulates the concatenation of f_1 and f_3, the sex path modulates a sex-specific projection of both. Because the two tasks compete for part of the shared feature space, decoupling lets each keep its own signal. A dedicated sex branch complements this: since the panoramic stream carries most sexual dimorphism (Results), f_1 is projected through a separate subnetwork to a 128-dimensional sex feature that reaches the sex head alongside the modulated representation. Sex prediction therefore does not depend on a representation shaped primarily for age.

The total loss combines the two objectives. A CORAL term (weight 0.5; the rank-consistent ordinal loss^77^) adds a ranking-based signal from the shared representation, and the sex weight of 3.0 compensates the gradient-magnitude imbalance between regression and classification so that age does not dominate optimisation. Alternative multi-task weighting schemes estimate these weights from task uncertainty^45,80^; fixed weights were used here instead (Eq. 4).

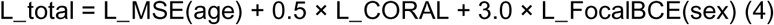

### Training Protocol and Computational Resources

Training protocol. Models were optimised with Adam^81^ at 1e-3 for the first 15 epochs with both backbones frozen, then at 1e-4 with all parameters trainable, halving the learning rate every 20 epochs (the two-phase schedule described in the Results). Mini-batches held 32 subjects drawn with a gender-balanced sampler. Augmentation combined a fixed jitter of 0.12, rotation of 5 degrees, feature dropout of 0.10, tooth dropout of 0.2, colour jitter and a tooth-context margin of 0.6, all held at mid-range values throughout training (fixed-mid augmentation in the Results); validation and test inputs were unaugmented. Training ran for a fixed 100-epoch budget with no early stopping, and the best validation checkpoint was retained for each head.

Computational resources. Models were trained on a single NVIDIA RTX 4080 SUPER (32 GB) with PyTorch 2.1.2, CUDA 12.1 and cuDNN 8.9. The final configuration took about 2.5 hours for 100 epochs; age metrics are taken from the best validation age epoch (51) and sex metrics from the best sex-accuracy epoch (46). The complete ablation study consumed roughly two days of GPU time.

### Evaluation Metrics and Statistical Analysis

#### Metrics

Age is evaluated by mean absolute error, MAE = (1/N) Σ_i |a_i − â_i|, with a_i and â_i the true and predicted ages in years, and sex by accuracy, SAcc = (1/N) Σ_i 1[g_i = ĝ_i]. Age metrics are reported at the epoch with the lowest validation MAE and sex metrics at the epoch with the highest validation sex accuracy (Results); ablation variants report both at their age checkpoint. Per-age-bin analysis stratifies seven intervals: 3–6, 6–8, 8–10, 10–12, 12–14, 14–16 and 16–18 years. Two trivial baselines bound the task: a majority-class sex predictor reaches 50% accuracy, and a constant-mean age predictor reaches an MAE equal to the mean absolute deviation of the dataset.

## Statistical analysis

Age errors are reported as MAE with standard deviation and sex as accuracy (SAcc). The main stream ablations (L1 only, L1+L3, L1+L2) were each trained with three seeds; the L1+L3 dual stream gives MAE = 0.52 ± 0.01 yr and sex accuracy = 85.8 ± 0.8% at 512 × 512 input. The deployed configuration (512 × 512 input, L1+L3 with GAT aggregation and FiLM fusion) reaches 0.51 yr and 85.2%; per-bin sample sizes accompany each value. All configurations were trained from random initialisation, and the four stream configurations were each run three times with independently drawn random seeds; because the seeds differ between configurations, runs are not matched across configurations and no run enters two comparisons. Configuration comparisons accordingly use unpaired tests that treat runs as replicates: two-sided Mann–Whitney U tests on absolute age errors and Fisher exact tests on sex classification, pooling the three runs of each configuration (n = 1,971 per arm), with Holm correction across age bins. Run-level Mann-Whitney U tests on the three seed means (n = 3 per arm) return P = 0.10 for each of the three main comparisons, the smallest two-sided value attainable at this sample size. The occlusion analysis (Supplementary Fig. 7) is the exception: the intervention is applied within the same images and each subject serves as its own control, so there the masked conditions are compared with the unoccluded reference using paired tests (two-sided Wilcoxon signed-rank on absolute age errors, exact McNemar on sex classification). The deployed 512 × 512 model has 46.4 M parameters and runs at 0.56 s per panoramic image with 0.55 GB of GPU memory at batch size 1 on a single RTX 4080 SUPER. Age error is governed by age rather than by the number of detected teeth, the partial correlation between tooth count and absolute error after conditioning on age being −0.12 (Supplementary Table 5). The 32- versus 52-class tooth comparison also uses paired tests, because both representations are evaluated on the same subjects.

### Input-utility criterion

For a candidate input X_new added to a specified retained set X_retained, the incremental utility for task t is ΔU_t(X_new | X_retained) = s_t(X_retained + X_new) − s_t(X_retained), where s_t is a task score oriented so that larger is better (s_age = −MAE; s_sex = accuracy). A positive value means improvement and a negative value degradation; because the scores carry different units they are never summed, so utility is a vector over tasks rather than one number. The same criterion can be stated as a relative change, (s_t(X_retained + X_new) - s_t(X_retained)) / |s_t(X_retained)|, which is the form used throughout the Results. The parent set is named with every comparison, and here X_retained = {L1} throughout. ΔU_t is therefore a first-order marginal at that parent and not order-independent: averaging over retained subsets gives the Shapley and SAGE families 78, 79, which three configurations do not permit us to estimate.

An input is retained only where ΔU_t is positive for at least one task and no task degrades beyond the seed-to-seed standard deviation of the compared configurations. Removal was implemented by retraining without the stream, which asks whether an input deserves to be kept; the tooth-level occlusion in the Results instead masks a structure at inference, which asks whether that structure is necessary for the prediction. ΔU_t is a property of the trained model rather than of the anatomy: a candidate that re-presents the same anatomy at another magnification, or with added annotations or relations, can change the increment, and the audit measures that effect instead of assuming it.

### Declaration

#### Data availability

The OPG images and the full subject-level labels cannot be made publicly available because of patient privacy regulations and institutional data-use agreements; qualified researchers may request access by contacting the corresponding author and submitting a data-use agreement approved by Nanfang Hospital. The public code repository (https://github.com/yingziyejiang/HMA- Net) provides the final model code, its parameter set and a small de-identified sample of the data; it does not contain the full subject-level labels.

#### Code availability

The final HMA-Net model code, the complete parameter set of the reported configuration, the trained tooth-detection weights are available at https://github.com/yingziyejiang/HMA-Net.

#### Ethics Approval

This study was approved by the Medical Ethics Committee of Nanfang Hospital, Southern Medical University (approval no. NFEC-2022-298). As a retrospective study of de-identified clinical images, the requirement for individual informed consent was waived. Names, hospital IDs and DICOM metadata containing protected health information were removed before transfer to the research server.

## Data Availability

The orthopantomograms and the subject-level labels cannot be made publicly available because of patient-privacy regulations and the institutional data-use agreement; qualified researchers may request access from the corresponding author under a data-use agreement approved by Nanfang Hospital. The model code, the reported configuration, the trained tooth-detection weights and a small de-identified sample (three cases with the expected input and label format) are publicly available at https://github.com/yingziyejiang/HMA-Net, frozen as the annotated tag v1.1-ncomms-submission (commit2b29cc224b8b4a37c69164c9c9eb631e96d015a7).

## Author contributions

**Zongze Wu:** Conceptualization, Methodology, Software, Investigation, Formal analysis, Writing – original draft. **Weijie Shan:** Conceptualization, Formal analysis, Validation, Visualization, Writing – original draft, Writing – review & editing. **Wujun Zhang:** Investigation, Data curation. **Zhenpei Kang:** Investigation, Data curation. **Benyu Yang:** Investigation, Data curation. **Zibin Liao:** Investigation, Data curation. **Zehang Huang:** Investigation, Data curation. **Yan Zhang:** Conceptualization, Project administration, Supervision, Writing – review & editing. **Xia Yue:** Conceptualization, Methodology, Formal analysis, Validation, Project administration, Supervision, Funding acquisition, Writing – review & editing.

All authors read and approved of the final manuscript.

## References

[1] Schmeling, A., Geserick, G., Reisinger, W. & Olze, A. Age estimation. Forensic Sci. Int. 165, 178–181 (2006).

[2] Willems, G. A review of the most commonly used dental age estimation techniques. J. Forensic Odontostomatol. 19, 9–17 (2001).

[3] Schmeling, A., Dettmeyer, R., Rudolf, E., Vieth, V. & Geserick, G. Forensic age estimation: methods, certainty, and the law. Dtsch Arztebl Int. 113, 44–50 (2016).

[4] Thevissen, P. W. et al. Human dental age estimation using third molar developmental stages. Forensic Sci. Int. 201, 106–111 (2010).

[5] Standing Committee of the National People’s Congress. Criminal Law Amendment of the People’s Republic of China (XI). http://www.npc.gov.cn/npc/c2/c30834/202012/t20201226_309442.html (2020).

[6] Stern, D., Payer, C., Giuliani, N. & Urschler, M. Automatic age estimation and majority age classification from multi-factorial MRI data. IEEE, J. Biomed Health Inform. 23, 1392–1403 (2018).

[7] Franklin, D., Flavel, A., Noble, J., Swift, L. & Karkhanis, S. Forensic age estimation in living individuals: methodological considerations in the context of medico-legal practice. Res. Rep. Forensic Med. Sci. 5, 53 (2015).

[8] Rozylo-Kalinowska, I. & Rozylo, T. K. Panoramic radiography in dentistry (Springer, 2021).

[9] Nolla, C. M. The development of the permanent teeth. J. Dent. Child. 27, 254–266 (1960).

[10] Liversidge, H. M., Herdeg, A. M. & Rosing, F. W. Dental age estimation of non-adults: a review of methods and principles. In Dental Anthropology 419–442 (Springer, 1998).

[11] Demirjian, A. Dental development: a measure of physical maturity. In Human Growth: A Comprehensive Treatise (eds Falkner, F. & Tanner, J. M.) 269–296 (Plenum Press, New York, 1986).

[12] Moorrees, C. F., Fanning, E. A. & Hunt, E. E. Age variation of formation stages for ten permanent teeth. J. Dent. Res. 42, 1490–1502 (1963).

[13] Liversidge, H. M. Variation in modern human dental development. In Patterns of Growth and Development in the Genus Homo (Cambridge University Press, 2003).

[14] Atas, I. Human gender prediction based on deep transfer learning from panoramic dental radiograph images. Trait. Signal. 39, 1585–1595 (2022).

[15] Park, S. J., Yang, S., Kim, J. M. & Kang, J. H. Automatic and robust estimation of sex and chronological age from panoramic radiographs using a multi-task deep learning network: a study on a South Korean population. Int. J. Legal Med. 138, 1741–1757 (2024).

[16] Franklin, D., O’Higgins, P., OXNARD, C. E. & Dadour, I. R. Discriminant function sexing of the mandible of Indigenous South Africans. Forensic Sci. Int. 179, 84.e1–84.e5 (2008).

[17] Capitaneanu, C., Willems, G. & Thevissen, P. A systematic review of odontological sex estimation methods. J. Forensic Odontostomatol. 35 (2017).

[18] Anderson, D. L. & Thompson, G. W. Interrelationships and sex differences of dental and skeletal measurements. J. Dent. Res. 52, 431–438 (1973).

[19] Demirjian, A., Goldstein, H. & Tanner, J. M. A new system of dental age assessment. Hum. Biol. 45, 211–227 (1973).

[20] Willems, G., Van Olmen, A., Spiessens, B. & Carels, C. Dental age estimation in Belgian children: Demirjian’s technique revisited. J. Forensic Sci. 46, 893–895 (2001).

[21] Cameriere, R., Ferrante, L. & Cingolani, M. Age estimation in children by measurement of open apices in teeth. Int. J. Legal Med. 120, 49–52 (2005).

[22] Esan, T. A., Yengopal, V. & Schepartz, L. A. The Demirjian versus the Willems method for dental age estimation in different populations: a meta-analysis of published studies. PLoS. One. 12, e0186682 (2017).

[23] Tomás, L. F., Mónico, L. D. S. M., Tomás, I., Varela-Patiño, P. & Martín-Biedma, B. J. The accuracy of estimating chronological age from Demirjian and Nolla methods in a Portuguese and Spanish sample. BMC Oral. Health. 14, 160 (2014).

[24] Merdietio Boedi, R., Angelakopoulos, N. & Franco, A. Evaluation of Demirjian staging method reproducibility and reliability for dental age estimation between eight forensic experts. Int. J. Legal Med. 139, 2883–2890 (2025).

[25] Campanella, G., Hanna, M. G. & Geneslaw, L. Clinical-grade computational pathology using weakly supervised deep learning on whole slide images. Nat. Med. 25, 1301–1309 (2019).

[26] Ahmedt-Aristizabal, D., Armin, M. A., Denman, S., Fookes, C. & Petersson, L. Graph-based deep learning for medical diagnosis and analysis: past, present and future. Sensors 21, 4758 (2021).

[27] Li, B. et al. Learning global and local features using graph neural networks for chest X-ray report generation. Signal Processing: Image Communication. 107, 116744 (2022).

[28] Vila-Blanco, N., Carreira, M. J., Varas-Quintana, P., Balsa-Castro, C. & Tomás, I. Deep neural networks for chronological age estimation from OPG images. IEEE Trans Med. Imaging. 39, 2374–2384 (2020).

[29] Yilmaz, E., Gorurgoz, C. & Kis, H. C. Forensic dental age estimation with deep learning: a modified Xception model for panoramic X-ray images. Forensic Sci. Med. Pathol. 21, 565–579 (2025).

[30] Hou, W. et al. Exploring effective DNN models for forensic age estimation based on panoramic radiograph images. Proc. IJCNN, 1–8 (2021).

[31] Upalananda, W., Prapayasatok, S., Lampang, S. N. & Chaichulee, S. The influence of integrating sex as a feature in deep learning-based dental age estimation using panoramic radiographs. Proc. BMEiCON., 1–5 (2023).

[32] Balel, Y., Sagtas, K. & Bulbul, H. N. Development and evaluation of a deep learning-based system for dental age estimation using the Demirjian method on panoramic radiographs. BMC Oral. Health. 25, 1172 (2025).

[33] Buyukcakir, B., Bertels, J., Claes, P. & Vandermeulen, D. OPG-based dental age estimation using a data-technical exploration of deep learning techniques. J. Forensic Sci. 69, 919–931 (2024).

[34] Wallraff, S., Vesal, S., Syben, C., Lutz, R. & Maier, A. Age estimation on panoramic dental X- ray images using deep learning. In Informatik aktuell 186–191 (Springer, 2021).

[35] Sivri, M. B., Taheri, S., Kırzıoğlu Ercan, R. G., Yağcı, Ü. & Golrizkhatami, Z. Dental age estimation: a comparative study of convolutional neural network and Demirjian’s method. J. Forensic Leg. Med. 103, 102679 (2024).

[36] Zhang, D., Yang, J., Du, S., Bu, W. & Guo, Y. C. An uncertainty-aware and sex-prior guided biological age estimation from orthopantomogram images. IEEE, J. Biomed Health Inform. 27, 4926–4937 (2023).

[37] Kokomoto, K. et al. Automatic dental age calculation from panoramic radiographs using deep learning: a two-stage approach with object detection and image classification. BMC Oral. Health. 24, 143 (2024).

[38] Mohammad, N., Muad, A. M., Ahmad, R. & Mohd Yusof, M. Y. P. Accuracy of advanced deep learning with TensorFlow and Keras for classifying teeth developmental stages in digital panoramic imaging. BMC Med. Imaging. 22, 66 (2022).

[39] Ong, S. H., Kim, H., Song, J. S., Shin, T. J. & Hyun, H. K. Fully automated deep learning approach to dental development assessment in panoramic radiographs. BMC Oral. Health. 24, 426 (2024).

[40] Kim, S., Lee, Y. H., Noh, Y. K., Park, F. C. & Auh, Q. S. Age-group determination of living individuals using first molar images based on artificial intelligence. Sci. Rep. 11, 1073 (2021).

[41] Zaborowicz, M., Zaborowicz, K., Biedziak, B. & Garbowski, T. Deep learning neural modelling as a precise method in the assessment of the chronological age of children and adolescents using tooth and bone parameters. Sensors 22, 637 (2022).

[42] Menze, B. H., Jakab, A. & Bauer, S. The multimodal brain tumor image segmentation benchmark (BRATS). IEEE Trans Med. Imaging. 34, 1993–2024 (2014).

[43] Huang, S. C., Pareek, A., Seyyedi, S., Banerjee, I. & Lungren, M. P. Fusion of medical imaging and electronic health records using deep learning: a systematic review and implementation guidelines. npj. Digit Med. 3, 136 (2020).

[44] Stahlschmidt, S. R., Ulfenborg, B. & Synnergren, J. Multimodal deep learning for biomedical data fusion: a review. Brief Bioinform. 23 (2021).

[45] Steyaert, S., Pizurica, M. & Nagaraj, D. Multimodal data fusion for cancer biomarker discovery with deep learning. Nat. Mach. Intell. 5, 351–362 (2023).

[46] Lipkova, J., Chen, R. J. & Chen, B. Artificial intelligence for multimodal data integration in oncology. Cancer Cell. 40, 1095–1110 (2022).

[47] Acosta, J. N., Falcone, G. J., Rajpurkar, P. & Topol, E. J. Multimodal biomedical AI. Nat. Med. 28, 1773–1784 (2022).

[48] Baltrušaitis, T., Ahuja, C. & Morency, L. P. Multimodal machine learning: a survey and taxonomy. IEEE Trans Pattern Anal. Mach. Intell. 41, 423–443 (2018).

[49] Wang, W., Tran, D. & Feiszli, M. What makes training multi-modal classification networks hard?. Proc. IEEE/CVF CVPR, 12692–12702 (2020).

[50] Huang, Y. et al. What makes multi-modal learning better than single (provably). Adv. Neural Inf. Process. Syst. 34 (2021).

[51] Huang, Y., Lin, J., Zhou, C., Yang, H. & Huang, L. Modality competition: what makes joint training of multi-modal network fail in deep learning? (provably). Preprint at 10.48550/arXiv.2203.12221 (2022).

[52] Nagrani, A. et al. Attention bottlenecks for multimodal fusion. Adv. Neural Inf. Process. Syst. 34 (2021).

[53] Matsuoka, Y., Ueno, Y. & Uehara, S. Deep-learning prostate cancer detection and segmentation on biparametric versus multiparametric magnetic resonance imaging. Int. J. Urol. 30, 1103–1111 (2023).

[54] Xu, J., Wei, H. & Qian, W. Incremental value of perivascular adipose tissue radiomics for non-contrast CT-based detection of aortic dissection. J. Imaging Inform Med. (2026).

[55] Yoon, J., Baek, N. & Yoo, R. E. Added value of dynamic contrast-enhanced MR imaging in deep learning-based prediction of local recurrence in grade 4 adult-type diffuse gliomas. Sci. Rep. 14, 2171 (2024).

[56] Brown, G., Pocock, A., Zhao, M. J. & Lujan, M. Conditional likelihood maximisation: a unifying framework for information theoretic feature selection. J. Mach. Learn Res. 13, 27–66 (2012).

[57] Liang, P. P., Zadeh, A. & Morency, L. P. Foundations and trends in multimodal machine learning: principles, challenges, and open questions. ACM Comput Surv. 56, 1–42 (2024).

[58] Jain, S. & Wallace, B. C. Attention is not explanation. Proc. NAACL-HLT, 3543–3556 (2019).

[59] Ying, R., Bourgeois, D., You, J., Zitnik, M. & Leskovec, J. GNNExplainer: generating explanations for graph neural networks. Adv. Neural Inf. Process Syst. 32 (2019).

[60] Yuan, H., Yu, H., Gui, S. & Ji, S. Explainability in graph neural networks: a taxonomic survey. IEEE Trans Pattern Anal. Mach. Intell. 45, 1–19 (2022).

[61] DeYoung, J., Jain, S. & Rajani, N. F. ERASER: a benchmark to evaluate rationalized NLP models. Proc. 58th. Annu. Meet. Assoc Comput Linguist., 4443–4458 (2020).

[62] Covert, I., Lundberg, S. M. & Lee, S. I. Explaining by removing: a unified framework for model explanation. J. Mach. Learn Res. 22, 1–90 (2021).

[63] Covert, I., Lundberg, S. M. & Lee, S. I. Understanding global feature contributions with additive importance measures. Adv. Neural Inf. Process Syst. 33 (2020).

[64] Bengio, Y., Louradour, J., Collobert, R. & Weston, J. Curriculum learning. Proc. ICML, 41–48 (2009).

[65] Loth, S. R. & Henneberg, M. Mandibular ramus flexure: a new morphologic indicator of sexual dimorphism in the human skeleton. Am. J. Phys. Anthropol. 99, 473–485 (1996).

66. Velickovic, P., et al. Graph attention networks. Proc. ICLR (2018).

[67] Pencina, M. J., D’Agostino, R. B. Sr., D’Agostino, R. B. Jr. & Vasan, R. S. Evaluating the added predictive ability of a new marker: From area under the ROC curve to reclassification and beyond. Stat. Med. 27, 157–172 (2007).

[68] Varma, S. & Simon, R. Bias in error estimation when using cross-validation for model selection. BMC Bioinformatics 7, 91 (2006).

[69] Dwork, C. et al. The reusable holdout: preserving validity in adaptive data analysis. Science 349, 636–638 (2015).

[70] Jocher, G., Chaurasia, A. & Qiu, J. Ultralytics YOLO8 (software), version 8.4.10. https://github.com/ultralytics/ultralytics (2023).

[71] He, K., Zhang, X., Ren, S. & Sun, J. Deep residual learning for image recognition. Proc. IEEE CVPR, 770–778 (2016).

[72] Luo, W., Li, Y., Urtasun, R. & Zemel, R. Understanding the effective receptive field in deep convolutional neural networks. Adv. Neural Inf. Process. Syst. 29 (2016).

[73] Wang, J., Sun, K. & Cheng, T. Deep high-resolution representation learning for visual recognition. IEEE Trans Pattern Anal. Mach. Intell. 43, 3349–3364 (2020).

[74] Goyal, A. & Bengio, Y. Inductive biases for deep learning of higher-level cognition. Proc. R. Soc. A. 478 (2022).

[75] Liu, Z. et al. A ConvNet for the 2020s. Proc. IEEE/CVF CVPR, 11966–11976 (2022).

[76] Perez, E., Strub, F., de Vries, H., Dumoulin, V. & Courville, A. FiLM: visual reasoning with a general conditioning layer. Proc. AAAI Conf. Artif Intell. 32 (2018).

[77] Cao, W., Mirjalili, V. & Raschka, S. Rank consistent ordinal regression for neural networks with application to age estimation. Pattern Recognit Lett. 140, 325–331 (2020).

[78] Lin, T. Y., Goyal, P., Girshick, R., He, K. & Dollar, P. Focal loss for dense object detection. Proc. IEEE ICCV, 2999–3007 (2017).

79. Standley, T., Zamir, A., Chen, D., Guibas, L., Malik, J. & Savarese, S. Which tasks should be learned together in multi-task learning? Preprint at 10.48550/arXiv.1905.07553 (2020).

[80] Kendall, A., Gal, Y. & Cipolla, R. Multi-task learning using uncertainty to weigh losses for scene geometry and semantics. Proc. IEEE CVPR, 7482–7491 (2018).

[81] Kingma, D. P. & Ba, J. Adam: a method for stochastic optimization. In Proc. International Conference on Learning Representations (ICLR) (2015).

