## Supplementary Information for "Removing Redundant Anatomical Inputs Improves Deep Learning-Based Forensic Dental Age and Sex Estimation"

### Supplementary Figures

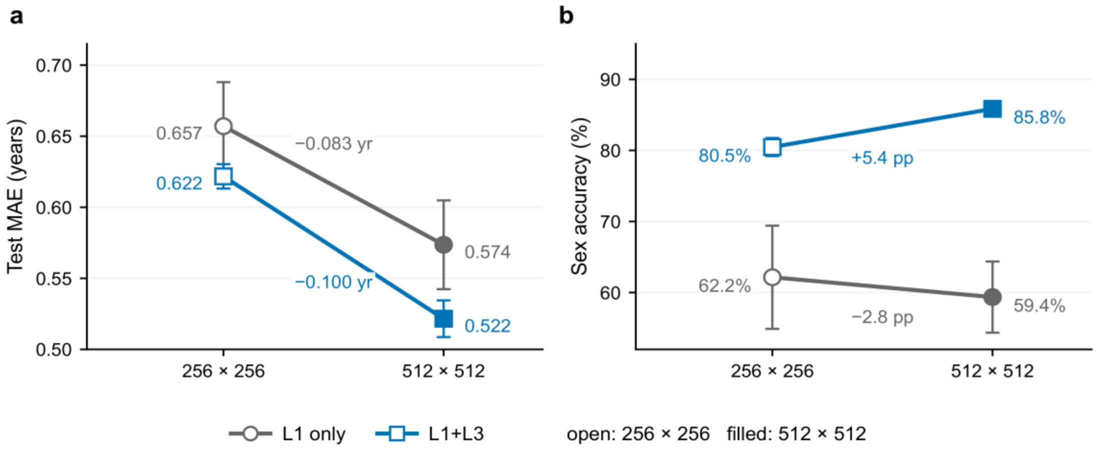


**Supplementary Fig. 1. Input-resolution ablation.**

Slope charts of (a) age MAE and (b) sex accuracy against input resolution for the panoramic-only and dual-stream configurations; open and filled markers denote 256 × 256 and 512 × 512, and error bars are SD over three seeds.

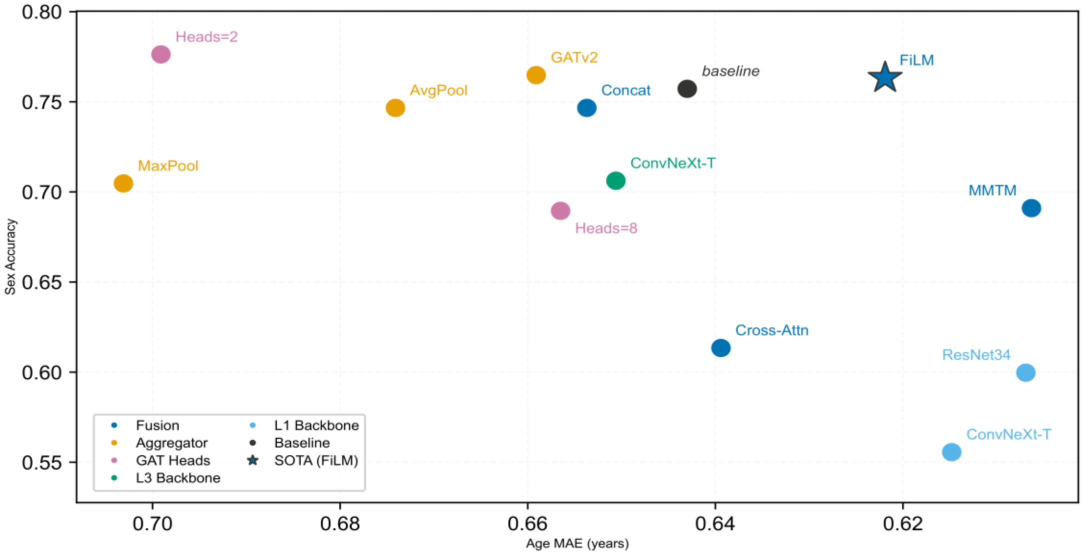


**Supplementary Fig. 2. Component ablation.**

Each point is one component substitution trained at the AGE checkpoint; the axes are age MAE (lower is better, axis reversed) and sex accuracy. Marker colour gives the substituted family (fusion, aggregation, GAT head count or stream backbone); the star marks the retained configuration.

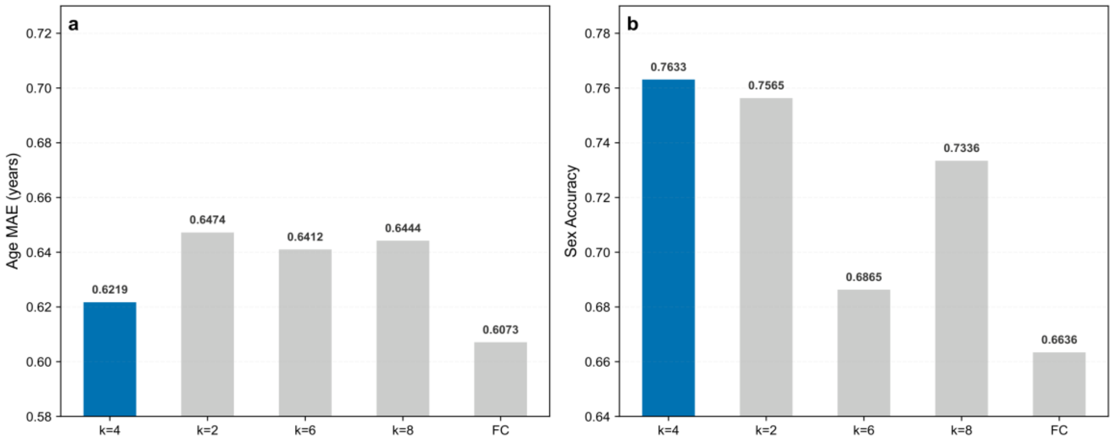


**Supplementary Fig. 3. Graph-topology ablation.**

(a) Age MAE and (b) sex accuracy for the neighbourhood degree k and a fully connected graph (FC); k = 4 is the retained topology.

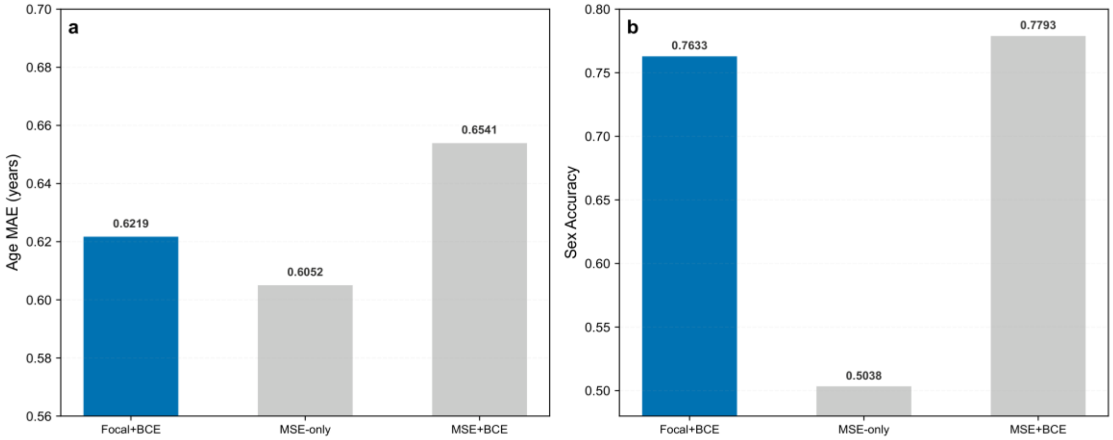


**Supplementary Fig. 4. Loss-composition ablation.**

(a) Age MAE and (b) sex accuracy. Removing the sex loss (MSE-only) collapses sex accuracy to chance; Focal BCE recovers it at limited age cost.

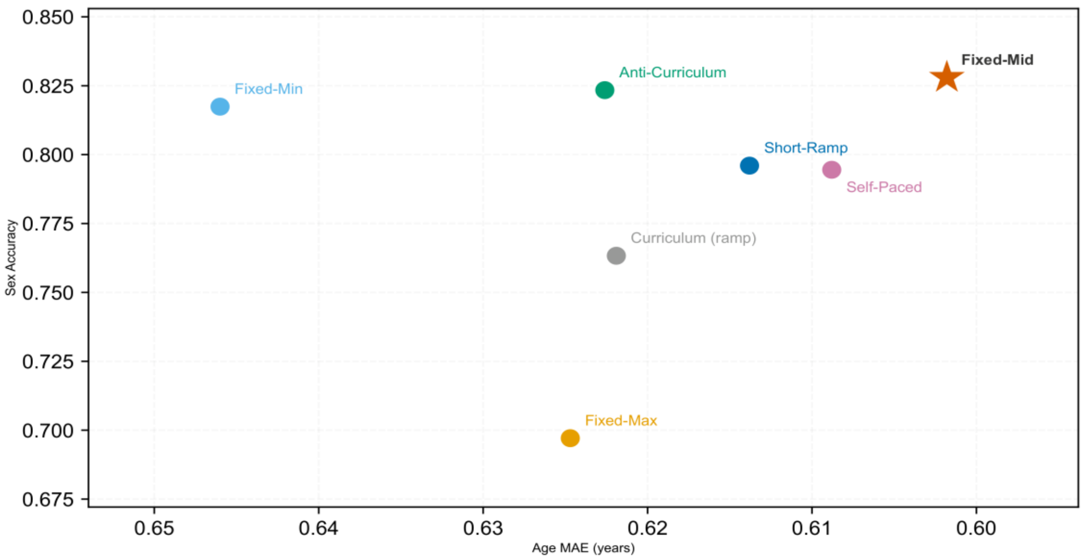


**Supplementary Fig. 5. Training-strategy ablation.**

Seven augmentation schedules scored by age MAE (lower is better, axis reversed) and sex accuracy at the AGE checkpoint; the star marks the retained schedule (fixed-mid).

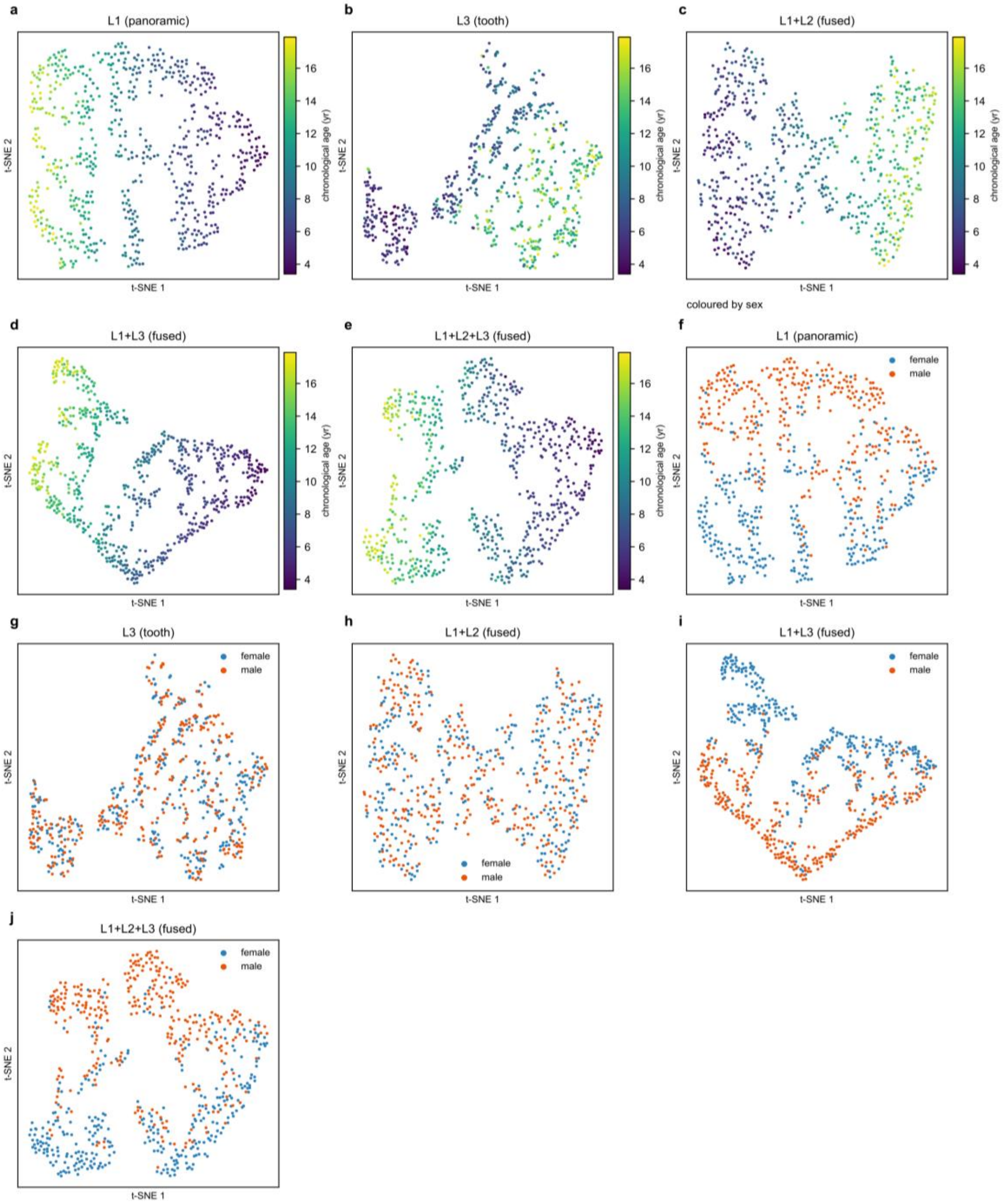


**Supplementary Fig. 6. t-SNE of the five stream configurations (panoramic, tooth, jaw-crop fusion, dual-stream fusion and three-stream fusion)**

(a–e) coloured by chronological age and (f–j) by sex; the five input configurations appear as pairs: (a, f) panoramic only, (b, g) tooth only, (c, h) jaw-crop fusion, (d, i) dual-stream fusion and (e, j) three-stream fusion.

33

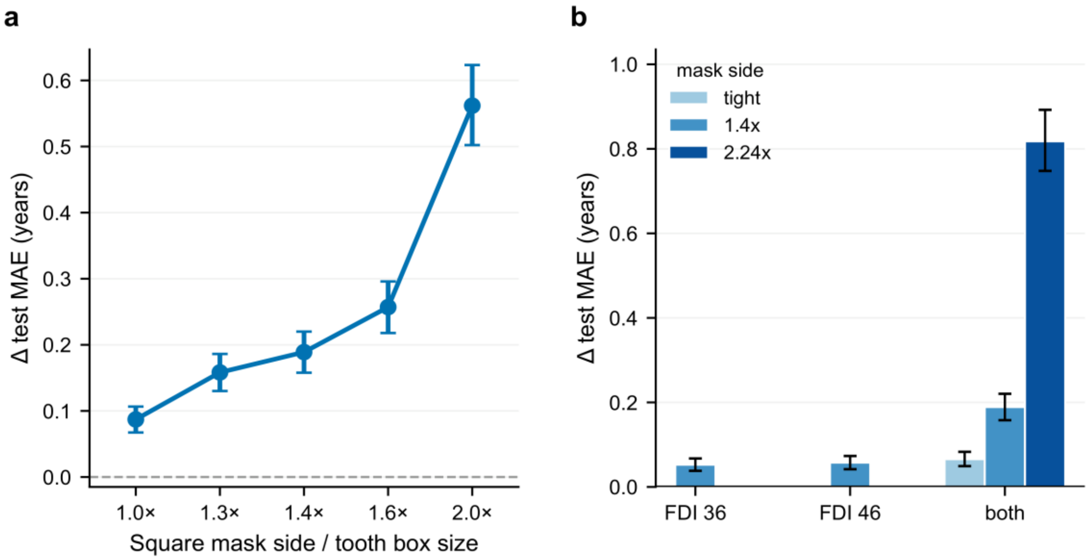


**Supplementary Fig. 7. Occlusion dose-response.**

(a) Paired change in age MAE and sex accuracy when both mandibular first molars (FDI 36 and 46) are occluded in both streams, as the square mask side grows from the tooth-box size to twice it. (b) Single-tooth versus both-teeth occlusion at three mask geometries (tight, 1.4-fold and 2.24-fold the tooth-box side). Masks zero the tooth ROI in the tooth stream and black the corresponding region of the panoramic input; all effects are paired mean differences with 95% bootstrap confidence intervals.

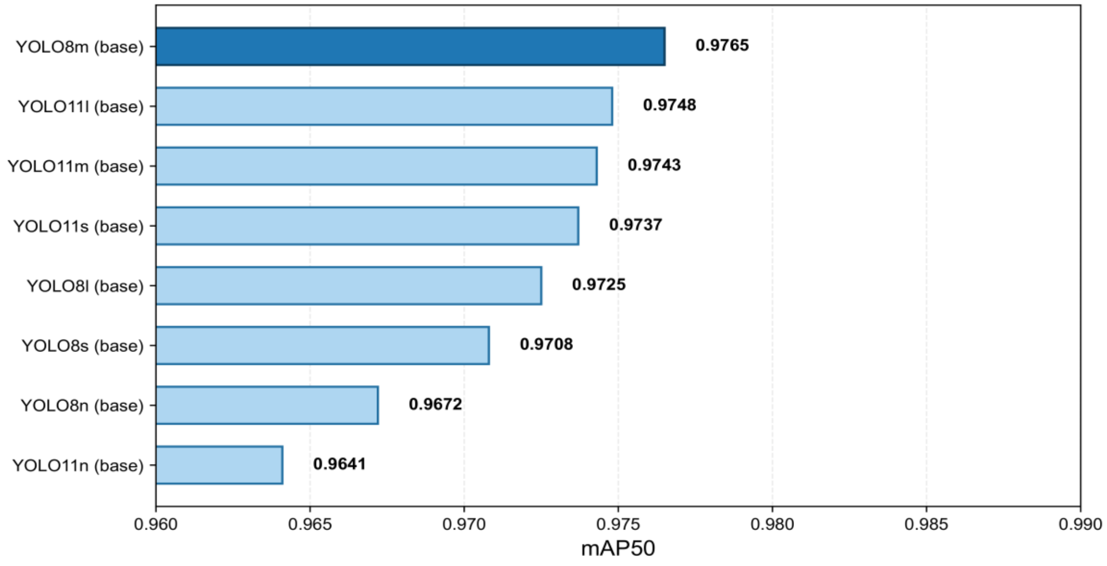

**Supplementary Fig. 8. Tooth-detector selection.**

Mean average precision at IoU 0.50 for eight YOLO OBB variants on 546 manually annotated panoramic images; YOLO8m-OBB was selected.

#### Supplementary Tables

**Supplementary Table 1.**

Sex-classification confusion matrix for HMA-Net (SAcc = 85.2%). Rows are true sex, columns predicted sex; cells are subject counts.

| **True \ Predicted** | **Female** | **Male** | **Total** |
|---|---|---|---|
| Female | 288 | 43 | 331 |
| Male | 54 | 272 | 326 |
| Total | 342 | 315 | 657 |

Per-class recall: female 0.870, male 0.834. Per-age-bin male/female recall: 3–6 yr 0.778/0.818; 6–8 yr 0.904/0.761; 8–10 yr 0.855/0.841; 10–12 yr 0.714/1.000; 12–14 yr 0.804/0.943; 14–16 yr 0.962/0.886; 16–18 yr 0.857/0.964.

**Supplementary Table 2.**

Threshold-level performance by sex. Sensitivity and specificity use the cut-off predicted age ≥ T; the last column is the misclassification rate among subjects whose true age lies within 0.5 yr of the threshold.

| Threshold (yr) | Sex | n | n ≥ T | AUC | Sensitivity | Specificity | Accuracy | Wrong call within ± 0.5 yr (%) |
|---|---|---|---|---|---|---|---|---|
| 12 | Female | 331 | 116 | 0.994 | 0.931 | 0.981 | 0.964 | 32.1 |
| 12 | Male | 326 | 93 | 0.992 | 0.914 | 0.974 | 0.957 | 37.5 |
| 14 | Female | 331 | 63 | 0.989 | 0.889 | 0.970 | 0.955 | 63.6 |
| 14 | Male | 326 | 47 | 0.997 | 0.936 | 0.996 | 0.988 | 19.0 |
| 16 | Female | 331 | 28 | 0.995 | 0.929 | 0.993 | 0.988 | 22.2 |
| 16 | Male | 326 | 21 | 0.994 | 1.000 | 0.980 | 0.982 | 13.6 |

**Supplementary Table 3.**

Comparison with published methods. N is the study cohort size as reported by each study; for HMA-Net it is the full collected cohort of 4,293 panoramic radiographs, of which 657 formed the independent test set used for all reported age and sex metrics. Columns give age range, MAE, sex accuracy (SAcc) and the evaluation setting; values for cited studies are those they reported, and cohorts, age ranges and protocols differ, so the values are not directly comparable.

| Method | Year | N | Age range | MAE | SAcc | Evaluation |
|---|---|---|---|---|---|---|
| HMA-Net (ours) | — | 4,293 | 3–18 | 0.51 | 0.852 | Independent test (n = 657) |
| Zhang et al. [60] | 2023 | 10,703 | 5–25 | 0.80 | NA | Internal test (regression) |
| Upalananda et al. [19] | 2023 | 1,734 | 8–23 | 0.70 | NA | Internal test (regression) |
| Sivri et al. [59] | 2024 | 5,898 | 4–17 | 0.73 | NA | Internal test (Demirjian stages) |
| Wallraff et al. [58] | 2021 | 14,000 | 11–20 | 1.08 | NA | Internal test |
| Buyukcakir et al. [51] | 2024 | 3,896 | <25 (adults & minors) | 0.56 | NA | Internal test (cross-validation) |

**Supplementary Table 4.**

Two-hop receptive field of the tooth graph as a function of k. Graphs were built as in the model (Euclidean k-nearest neighbours on bounding-box centres) from the detected boxes of all 4,299 labelled OPGs; values are the mean two-hop reachable set per node.

| k | Mean teeth reached | SD | Median | Quadrant equivalent | Samples with >= 8 teeth |
|---|---|---|---|---|---|
| 2 | 4.58 | 0.20 | 4.64 | 0.57 | 0.0% |
| 3 | 6.56 | 0.25 | 6.61 | 0.82 | 0.0% |
| 4 | 8.49 | 0.32 | 8.50 | 1.06 | 94.9% |
| 5 | 10.36 | 0.47 | 10.39 | 1.30 | 99.9% |
| 6 | 12.28 | 0.66 | 12.36 | 1.54 | 100.0% |
| 7 | 14.33 | 0.85 | 14.36 | 1.79 | 100.0% |
| 8 | 16.43 | 1.19 | 16.44 | 2.05 | 100.0% |

**Supplementary Table 5.**

Age error and sex accuracy by number of detected teeth (deployed model). Teeth detected is the number of permanent teeth returned by the detector for that image, counting one box per FDI number. The unadjusted tooth-count trend is an age effect: conditioning on age, the partial correlation between tooth count and absolute error is −0.12.

| Teeth detected | n | Age MAE (yr) | Sex accuracy | Mean age (yr) |
|---|---|---|---|---|
| 10–25 | 126 | 0.377 | 0.849 | 6.26 |
| 26–27 | 132 | 0.505 | 0.803 | 7.70 |
| 28–30 | 229 | 0.497 | 0.869 | 9.99 |
| 31–32 | 170 | 0.622 | 0.871 | 13.48 |
| All | 657 | 0.508 | 0.852 | 9.72 |
